# Sensitivity to climate change is widespread across zoonotic diseases

**DOI:** 10.1101/2024.11.18.24317483

**Authors:** Artur Trebski, Lewis Gourlay, Rory Gibb, Natalie Imirzian, David W. Redding

**Author notes:** These authors contributed equally (Artur Trebski, Lewis Gourlay).

## Abstract

Climate change is expected to exacerbate infectious diseases, yet the climate sensitivity of zoonotic diseases (driven by spillover from animal reservoirs) is markedly understudied compared to vector-borne and water-borne infections. To address this gap, we conducted a global systematic review and quantitative synthesis to identify relationships between climatic indicators (temperature, precipitation, humidity) and zoonotic disease risk metrics worldwide. We identified 185 studies from 55 countries, describing 547 measures across 51 diseases, with most studies testing linear (n=166) rather than nonlinear (n=23) relationships. We found evidence of climate sensitivity across diverse zoonotic diseases (significant non-zero relationships in 64.3% of temperature effects, 49.8% of precipitation effects, and 48.9% of humidity effects), but with broad variation in direction and strength. Positive effects of temperature and rainfall on disease risk were more common than negative effects (39.1% vs. 25.2% and 30.5% vs. 19.2% of all records, respectively). These studies were predominantly located in areas expected to have substantial increases in annual mean temperature (>1.5°C in 93% of studies) and rainfall (>25 mm in 46% of studies) by 2041–2070. Notably, the most consistent relationship was between temperature and vector-borne zoonoses (50% of Positive effects, mean Hedge’s g = 0.31). Overall, our analyses provide evidence that climate sensitivity is common across zoonoses, likely leading to substantial yet complex effects of future climate change on zoonotic burden. Finally, we highlight the need for future studies to use biologically appropriate models, rigorous space-time controls, consider causal perspectives and address taxonomic and geographic biases to allow a robust consensus of climate-risk relationships to emerge.

**Significance statement:** Understanding how climate change affects zoonotic diseases—those transmitted from animals to humans—is crucial for public health planning yet remains underexplored. Our global analysis of 185 studies covering 51 zoonotic diseases reveals widespread climate sensitivity among these diseases. Climatic factors, particularly temperature, are often linked to increased disease risk, especially for vector-borne diseases transmitted by arthropods. With many regions projected to experience significant warming, climate change may exacerbate zoonotic disease burden. However, few studies have considered nonlinear effects, and the variation in responses both within and across diseases indicates complex dynamics that require biologically informed research methods. These findings underscore the urgent need for improved research approaches to better predict and manage future disease risks in a changing climate.

## Introduction

The rapid increase in the emergence of zoonotic diseases and diseases of zoonotic origin, such as Zika, Ebola and COVID-19, presents significant threats to global economies, public health, and social stability (1). The processes that drive pathogen spillover operate at the nexus of environmental change, socioeconomic structure, and public health (2). As global changes—including climate change, urbanisation, and land-use transformation—reshape human-environment interfaces, the risk of novel patterns of zoonotic disease transmission rises (3–5). Among these drivers, climate change stands out as a particularly important yet still poorly understood factor (6), necessitating additional research into how it shapes zoonotic disease burden across various systems.

Despite most recent pandemics being zoonotic in origin, most of the previous research on climate change as a moderator of pathogen spillover has focused on high burden, vector-borne diseases (7–10). Moreover, biases in reporting efforts for neglected or emerging zoonoses — particularly in under-resourced settings — add to the challenge of establishing baseline evidence (11, 12). Addressing these knowledge gaps is essential for developing realistic projections of how climate change may impact the global burden of zoonotic diseases (13). A better understanding of these dynamics is also crucial for developing global longitudinal monitoring programs, informing current research priorities, and helping identify potential trends in future zoonotic risk (3, 8, 14).

To extrapolate zoonotic disease risk into the future, it is vital to identify trends in the local-scale climate sensitivity of pathogens. Short-term variation in key climatic factors, such as temperature, precipitation, and humidity, can alter disease transmission through multiple mechanisms, such as modified contact rates between hosts, impacts on vector thermal responses, and changes to host population dynamics (3, 15–20). For example, seasonal changes in temperature and precipitation drive both vector population sizes and thermal suitability for mosquito-borne disease transmission (e.g., dengue, malaria, Zika) (9, 21–23). Similarly, rodent population cycles are strongly influenced by seasonal and multi-year precipitation, consequently impacting the spillover dynamics of zoonotic pathogens such as hantaviruses, Lassa virus and leptospirosis (2, 24–27).

Given the evidence of climate sensitivity in zoonotic diseases, transmission risk is likely to be impacted by climate change. Long-term changes to the climate may drive large-scale movement of key hosts and vectors, modify land-use patterns, and alter basic physiological responses with consequences for disease susceptibility (3, 18, 28–30). This may both exacerbate and suppress different elements of transmission, creating potentially complex effects that vary across systems (20). Improving our knowledge of the response of reservoir hosts to climate variability will help disentangle potential impacts of climate change on disease risk. While many mosquito-borne diseases are well-studied in relation to climate variability (22), the climate sensitivity of zoonoses more broadly is poorly defined, despite representing an important dimension of public health risk.

Recent reviews and analyses have addressed some key questions in this area. Many studies identified increasing disease risk stemming from climate change (5, 19, 20, 31), yet they reveal mixed findings on the consistency of these responses, with some showing high degree of context-dependent effects. Notably, a gap remains in research specifically addressing the sensitivity of diverse zoonotic disease systems to different climate factors. By synthesising globally distributed empirical studies on various climate metrics and disease risk measures, this study aims to uncover broader patterns of climate sensitivity unique to zoonoses.

Here, we conduct a comprehensive analysis of primary studies that evaluate the empirical relationships between climatic parameters (i.e., temperature, precipitation, and humidity) and measures of zoonotic risk (i.e., abundance, seroprevalence, number of cases, and incidence) to assess the extent of climate sensitivity across zoonotic disease systems. Specifically, our study aims to: (1) identify which regions and zoonotic diseases are over– or underrepresented in climate sensitivity research; (2) determine whether any consistent patterns emerge in climate sensitivity across disease types, regions, types of vectors, and hosts; (3) evaluate if the methods and metrics used in source studies are appropriate to detect such trends; and (4) assess whether climate sensitivity differs by transmission pathways.

## Results

### Literature search results

The search criteria (Table S1) yielded 13,468 article titles, which were narrowed down through a four-step screening process (Fig. S1) to 185 (1.3% of total titles) independent, empirical studies that could be included in the final dataset. These studies, representing 51 diseases (Table S2) from 55 countries, provided 547 statistics quantifying the relationship between climate variables and various measures of zoonotic disease risk. Temperature was the most commonly studied variable (43%, n=236), followed by precipitation (40%, n=218), and humidity (17%, n=93). The studies had close to global coverage, with areas of high sampling in South and East Asia and Europe, and more limited sampling in Central and North Asia and Eastern Africa (Fig. 1A). Hantaviruses were the most frequently studied group of pathogens (25.1%, n=49), followed by arboviruses (17.9%, n=35) and *Leptospira* spp. (12.3%, n=24) (see Fig. 1B).

**Figure 1:**
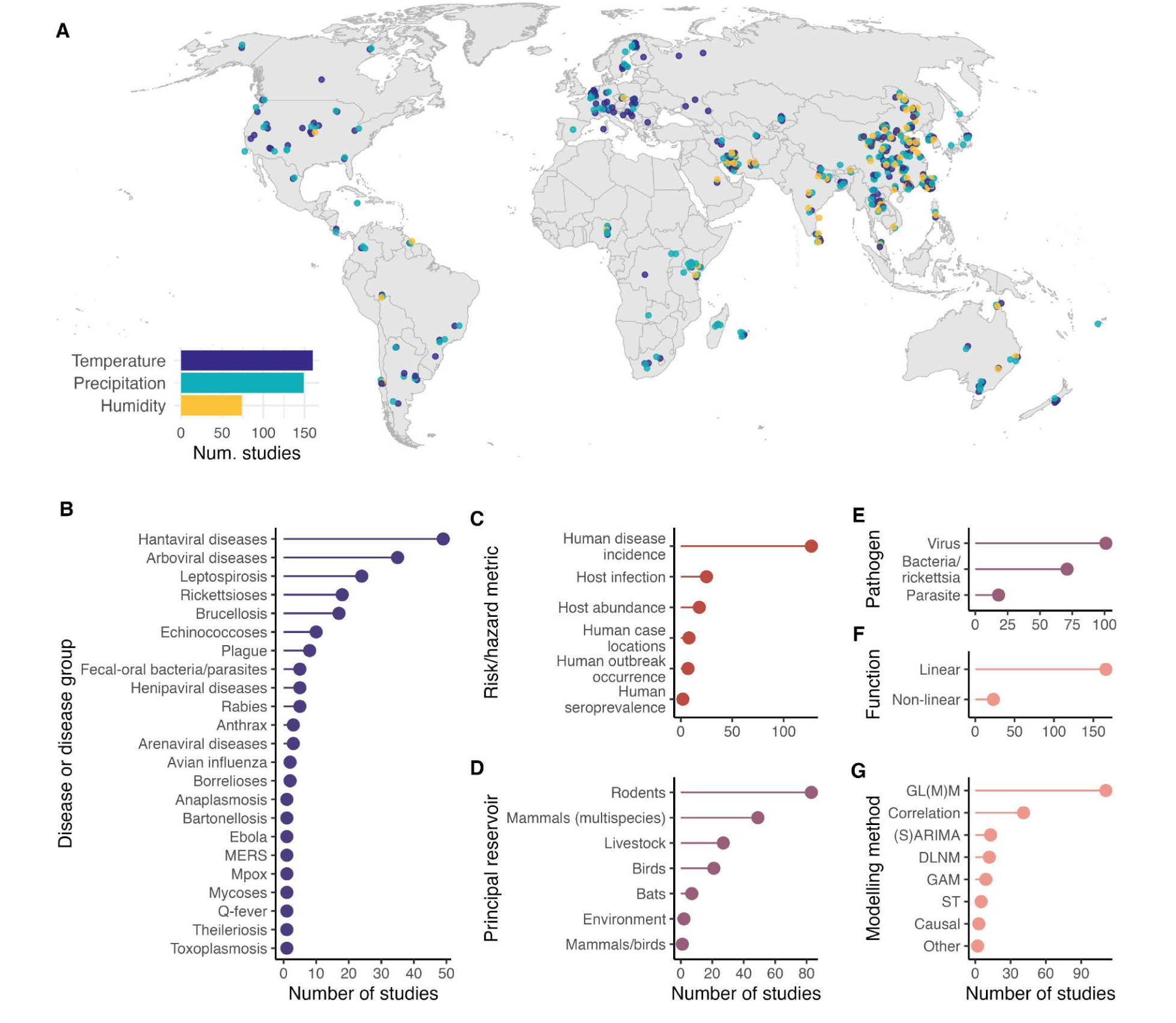
Meta-analysis database of climatic driver effects on zoonotic disease transmission. Map shows the full database of extracted climate effects (n=547 effects, 185 studies) summarised geographically, with point colour representing climatic driver, and location representing the specified lat-lon or nearest named locality of the study. Studies without locality information were geolocated to the country centroid. Inset barplot shows the number of studies reporting effects of temperature, precipitation or humidity on zoonotic diseases. Subplots show the database broken down by key variables: disease or broad disease grouping; the risk or hazard metric being tested in the study; the principal reservoir host(s) of the study’s focal disease; the broad pathogen type; whether the study reported linear or nonlinear inferred effects; and the broad type of modelling method used. Method abbreviations: GL(M)M, generalised linear (mixed effects) model; (S)ARIMA, (seasonal) autoregressive integrated moving average model; DLNM, distributed-lag nonlinear model; GAM, generalised additive model; ST, spatiotemporal statistical model; Causal, an explicitly causal inference-based model; Other, descriptive or basic frequentist statistics (e.g. Chi-square test).

### Zoonotic risks are generally higher in warm and wet conditions

In general, climate factors influence zoonotic disease risk to some extent, according to the majority of measures assessed (n = 547; 72% significant at α=0.05, 42% at α=0.01, 22% at α=0.001). Out of the 185 source studies, 137 (74%) reported more than one measure of zoonotic risk with an associated p-value. Among these 137 studies with secondary reporting, 131 (96%) had at least one measure of zoonotic disease risk significant at the α=0.05 level, while 103 (75%) reported two or more significant measures of zoonotic risk at the α=0.05 level.

There were significantly more positive relationships between climatic factors and a measure of risk (1.7 times more) reported than negative relationships (χ̅² = 21.35, df = 1, 95% CI = 20.82 – 21.89, p = 0.001, from 1000 bootstraps of 80% of the data). The proportions seen within the overall dataset were robust to dropping most major regions, disease groups, pathogen types, statistical methods, and hosts from the calculations (Table S3). When dividing the dataset into vectored and non-vectored diseases, only the subset containing vectored diseases showed significantly more positive relationships (n = 101) than negative relationships (n = 34) (Non-vectored diseases: χ̅² = 4.65, df=1, p= 0.164; Vectored diseases: χ̅² = 27.06, df=1, p= 0.0002; Table S3). However, non-vectored diseases also exhibited a substantial number of positive relationships (positive: n = 146; negative: n = 113).

### Reported climate-risk relationships are highly variable in size and direction both between and within different zoonotic disease systems

After standardising the measures of disease risk by transforming them into Hedge’s g, studies still reported an effect of climatic factors on zoonotic disease risk more often than no effect (56% Non-zero effect sizes; Fig. 2). However, when splitting the dataset into the three climatic factors separately, only temperature reported a non-zero effect more often than no effect (64.3% Non-zero effect sizes), with less than half of precipitation (49.8%; n=107) or humidity (48.9%; n=47) observations reporting a non-zero effect on disease risk.

**Figure 2.**
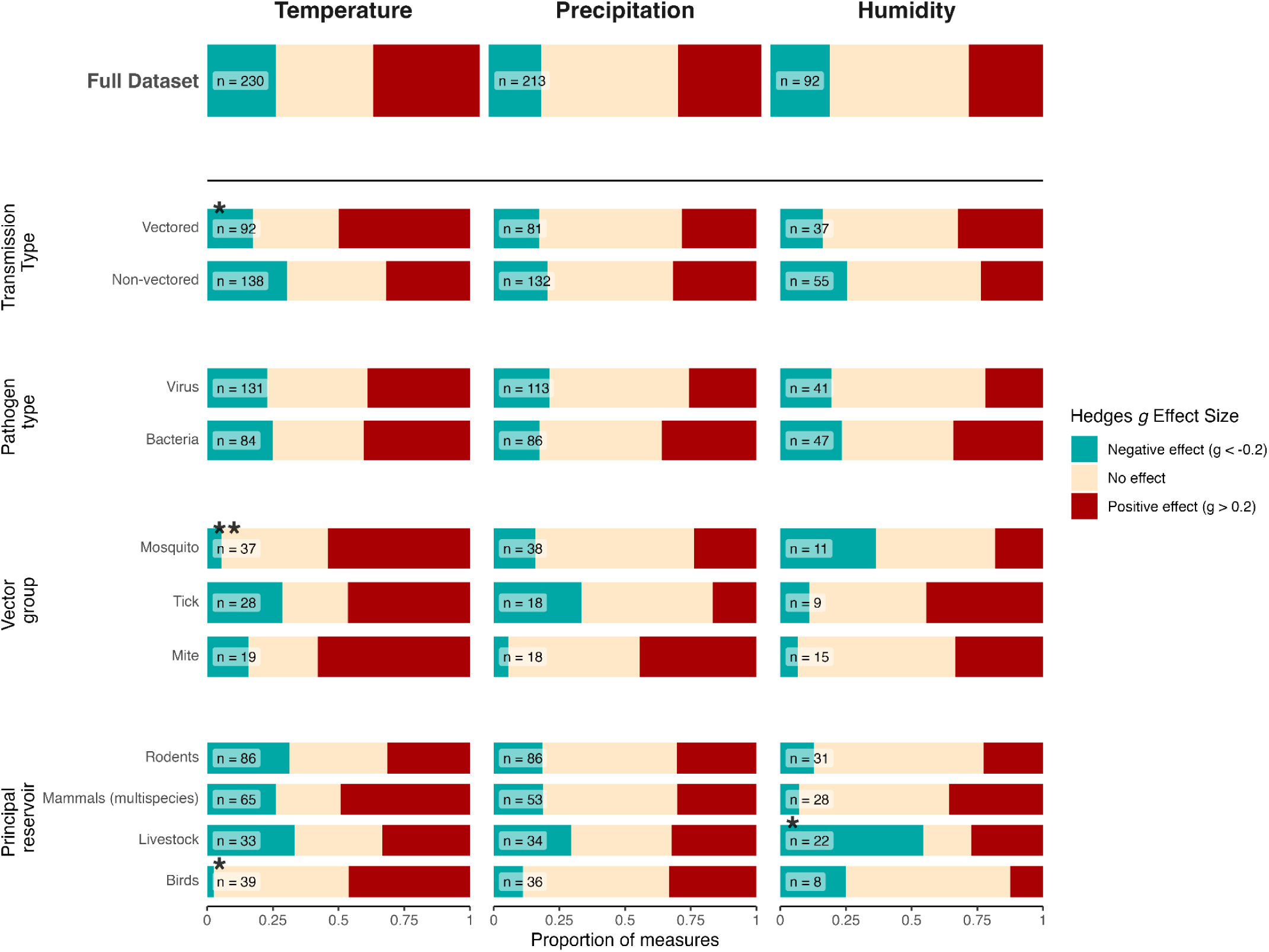
Direction of climate effects across key disease variables. Each column represents the climate driver studied, with the x-axis indicating the proportion of study measures falling into effect categories from a Negative effect (blue) to No effect (cream) to a Positive effect (red). The rows separate the data into the key variables identified in Figure 1: the main transmission mode (vectored via a biting vector from reservoir host to human, or non-vectored, with no involvement of a vector between reservoir host and human); the broad pathogen type; for the vector-borne diseases, the broad animal group of the vector; and the principal reservoir host(s) of the study’s focal disease. Groupings were included if the category had greater than 15 effect size measures. The number of measures included in each grouping is indicated next to the bars. Distributions that were significantly different from the full dataset are marked with asterisks above the sample size (* = p < 0.05; ** p < 0.01).

Overall, temperature appears to be the most consistent in terms of the direction of climate effects on disease risk (39.1% Positive effects, 35.7% No effects, 25.2% Negative effects). When comparing the three climatic factors, temperature showed the highest proportion of Positive effects on measures of disease risk, with 39.1% (n=90) of effects having a value of Hedge’s g greater than 0.2 (Fig. 2), with precipitation and humidity having more balanced Positive and Negative effects (Precipitation: 30.5% Positive and 19.2% Negative effects; Humidity: 27.2% Positive and 21.7% Negative effects). Furthermore, when grouping the effect sizes by disease and climatic factor, we observe a greater number of Positive effects in certain diseases, such as temperature impacts on West Nile Virus and Japanese encephalitis risk (Fig. 3A), and precipitation impacts on Japanese encephalitis, Scrub typhus, and Leptospirosis risk (Fig. 3B).

**Figure 3.**
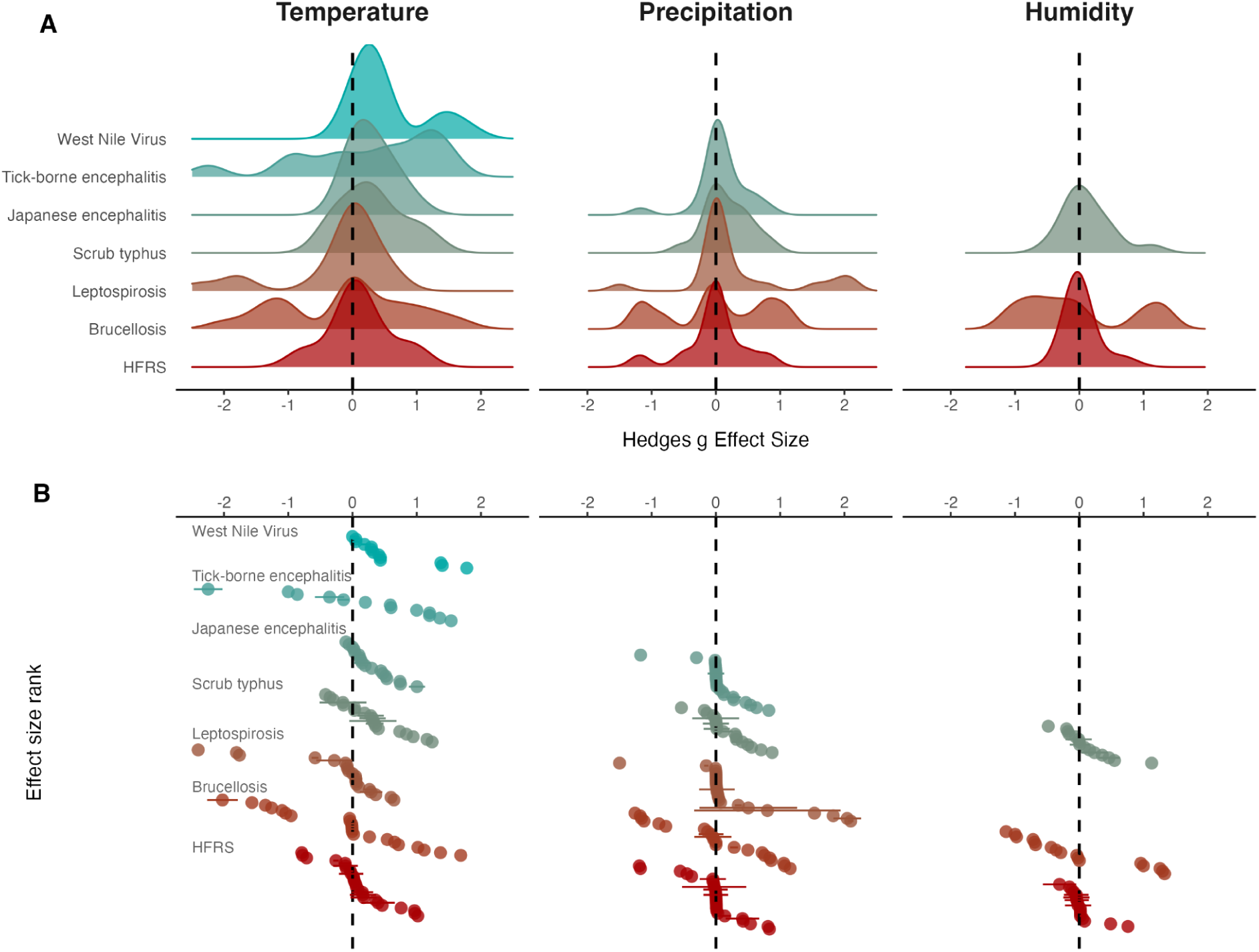
Comparison of effect size distribution across specific diseases. **(A)** Density plots showing the distribution of Hedges *g* values calculated from studies measuring disease risk in response to three climate variables: temperature, precipitation, and humidity (bottom row). The bluish plots represent vectored diseases (West Nile Virus, Tick-borne encephalitis, Japanese encephalitis and Scrub typhus) and the reddish density plots represent non-vectored diseases (Leptospirosis, Brucellosis and HFRS – Haemorrhagic fever with renal syndrome), for which more than ten data points were available under each climate variable. Extreme values (Hedges *g* values less than -2.5 or greater than 2.5) were excluded from the plot. **(B)** A scatterplot displaying the effect size values contributing to the distributions; each point represents the effect size calculated from a study, and points are ordered on the y-axis from lowest to highest values. Lines show the 95% confidence intervals around the values where the data permitted such calculations.

### Vector-borne zoonoses show the strongest and most consistent evidence for climate-sensitivity

The effect size distributions for vector-borne diseases, diseases vectored by mosquitoes, and diseases with birds as principal reservoir significantly differed from the overall effect size distribution for temperature, suggesting a positive influence of temperature on disease risk among these groups (Anderson-Darling Test; vector borne disease: AD = 2.33; p=0.047; mosquito vectored disease: AD = 3.63; p=0.008; birds as principal reservoir: AD = 3.1; p=0.025). When the subsets were compared to the full distributions with the subset removed, a few more categories were significantly different: non-vectored diseases and diseases located in Iran significantly differed from the overall temperature distribution (Table S4). Importantly, vector-borne zoonotic diseases had significantly more Positive effects (mean=0.31; sd=0.89; n=92) reported for temperature when compared to solely animal-hosted diseases (mean=-0.02; sd=1.01; n=138) (Two-sample Kolmogorov-Smirnov test; D=0.26; p = 0.001; Fig. 4, Table S5). While the trends for non-vectored diseases were less pronounced, 32% of temperature effect sizes for these diseases were Positive (compared to 50% for vector-borne diseases), indicating that temperature increases may still elevate the risk for a range of non-vectored zoonotic diseases.

**Figure 4.**
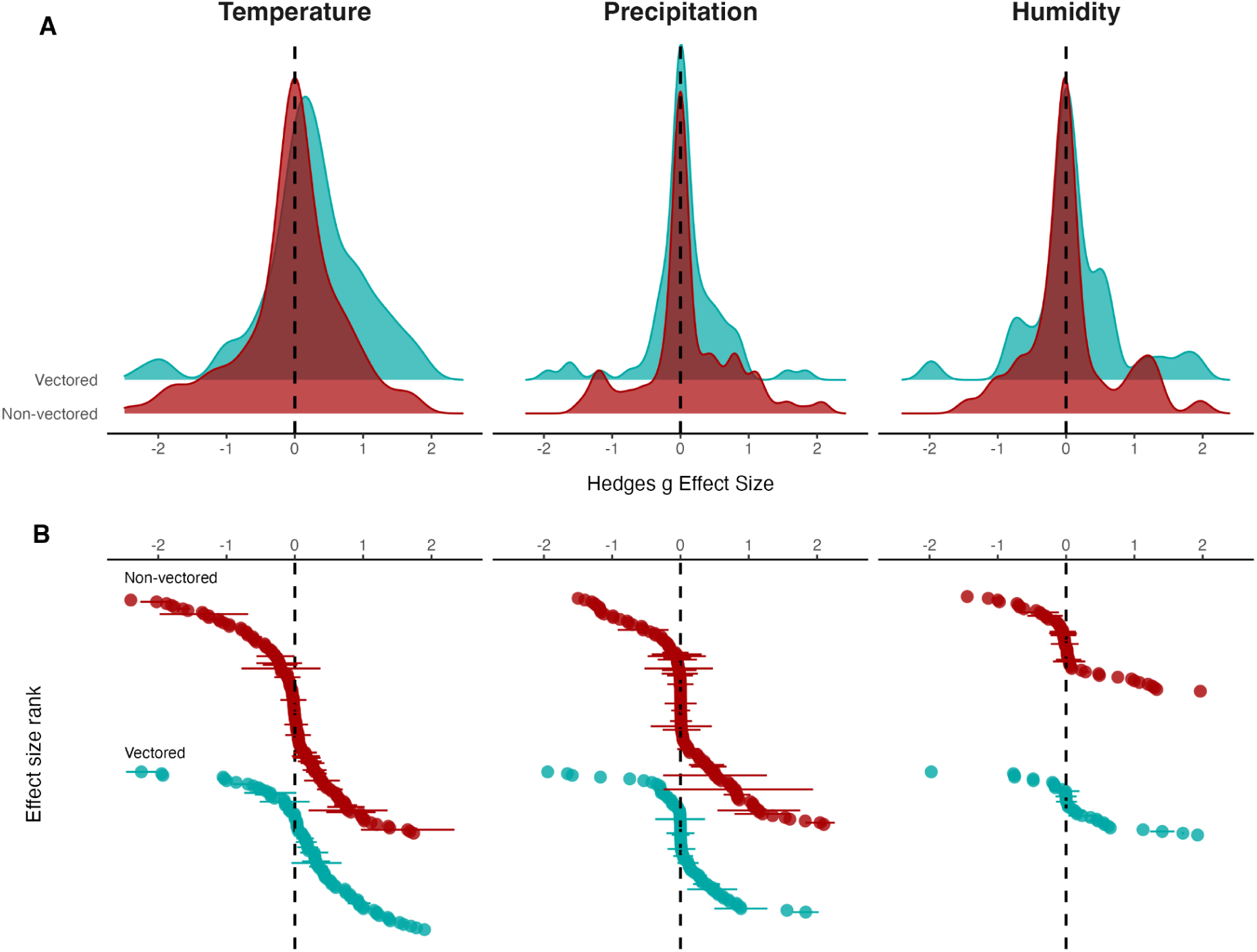
Comparison of effect size distribution between vectored and non-vectored diseases. **(A)** Density plots showing the distribution of Hedges *g* values calculated from studies measuring vector-borne and non vector-borne zoonotic disease risk in response to three climate variables: temperature, precipitation, and humidity (bottom row). The blue density plot represents vectored disease and the red density plot represents non-vectored disease. Extreme values (Hedges *g* values less than -2.5 or greater than 2.5) were excluded from the plot. **(B)** A scatterplot displaying the effect size values contributing to the distributions; each point represents the effect size calculated from a study, and points are ordered on the y-axis from lowest to highest values. Lines show the 95% confidence intervals around the values where the data permitted such calculations

The effect size distributions for both precipitation and humidity centred around zero, with no significant differences observed between the distributions of vectored and non-vectored subsets (Two-sample Kolmogorov-Smirnov test for precipitation: D=0.09; p = 0.445; and humidity: D=0.23; p = 0.079, Fig. 2 & Fig. 4, Table S5). We note the distribution of effect sizes for precipitation had visually larger tails for non-vectored diseases compared to vectored diseases (Fig. 4B), but the available data was too limited to perform robust statistical tests. Moreover, while the majority of observations on the impacts of humidity on disease risk showed no significant effects, a subset of studies on diseases with livestock as the principal reservoir and studies in Iran did exhibit significantly different distributions of effect sizes for humidity (Anderson-Darling Test; Livestock: AD = 2.74; p=0.036; Iran: AD = 3.43; p=0.016; Fig. 2, Table S6). When the distribution of effect sizes within specific subsets were compared to the full distributions with the subset removed, diseases with a livestock principal reservoir differed from the overall precipitation distribution (Table S7).

### Future climatological changes at study locations may favour increases in zoonotic transmission risk

We identified that many study sites associated with climate-sensitive diseases (those with Hedge’s g > 0.2 or < -0.2) are located in areas where future climatologies consistently predict substantial changes in temperature (BIO1, mean annual air temperature) or precipitation (BIO12, annual precipitation amount). These predictions, based on the CHELSA V2.1 CMIP6 dataset, were assessed across three SSP scenarios (SSP1-RCP2.6, SSP3-RCP7.0, SSP5-RCP8.5) and five GCMs for the period 2041–2070, compared to baseline conditions (1981–2010). Climate shifts were categorised using thresholds for temperature (1°C, 1.5°C, and 2°C) and precipitation changes (±25 mm, ±50 mm, ±100 mm), identifying the most congruent range of predictions for each location at each threshold.

A majority of study sites for which we identified Positive disease risk effects are located in areas expected to see substantial increase in temperature (100% of studies were in sites predicting >1°C increase, 94% above 1.5°C and 64% above 2°C; Fig. S6). Negative effect sizes observed a similar relationship, with most sites also seeing future temperature increases, meaning there was no association between effect size categories (Positive/Negative) and temperature change (Table S8).

The projected changes in precipitation across sites revealed more variability compared to temperature shifts, though again with no clear association between precipitation climate sensitivity categories (Positive/Negative) and predicted changes in annual precipitation (Table S8). For sites with Positive precipitation effect sizes, 46% saw an above +25mm increase in annual precipitation, 35% above

+50mm and 22% above 100mm (Fig. S6). Substantial decreases in precipitation were rarely predicted, meaning few sites with Negative precipitation sensitivity were located in areas expected to see lower precipitation (5% of study sites showing Negative effects were predicted a 25mm reduction, 0% a 50mm or 100mm reduction; Fig. S6). For both Positive and Negative rainfall effects, many sites are expected to see intermediate shifts in precipitation.

### Data biases and methodological inconsistencies across studies hinder a clear synthesis of climate-sensitivity in zoonoses

Several inconsistencies and methodological limitations were identified within the collated dataset and source studies. While Hedge’s *g* was able to be calculated for a total of 535 measures (98% of the extracted measures), the confidence intervals around Hedge’s g were able to be calculated for only 253 effect sizes, due to limited reporting of statistics within source studies and a lack of clearly reported sample sizes, standard errors or confidence intervals.

The dataset analysed contains considerable diversity in both the number of data points per disease and the statistical methods employed across studies. For example, certain diseases were well-represented including haemorrhagic fever with renal syndrome (n=28 studies) and leptospirosis (n=23), while eight diseases (anaplasmosis, bartonellosis, Ebola, MERS, mPox, Q-fever, theileriosis, and toxoplasmosis) were represented by just one study each. However, even among well represented diseases, effects were highly variable in both magnitude and direction (regardless of transmission type), further limiting the identification of clear patterns of climate sensitivity (Fig. 3).

In terms of methodology, the collated dataset represented 82 specific statistical methods used in the source studies, many of which lacked consistent and detailed reporting of methods used. The vast majority applied linear regression or time-series modelling approaches, with fewer studies applying spatially-explicit, nonlinear or explicitly causal inference-based modelling methodologies (Fig. 1). The heavy reliance on linear regression and time-series models may oversimplify the complexities of climate-disease relationships. Strikingly, the use of non-linear models was relatively rare, with only 12.1% of studies investigating non-linear relationships between climatic variables and measures of zoonotic disease risk (Fig. 1F).

There was some degree of publication bias inferred from funnel plots showing standard errors against respective effect sizes (Fig. S3). Egger’s regression tests for funnel plot asymmetry showed significant asymmetry in the overall dataset (z = 2.62, p = 0.009) and precipitation data (z = 2.71, p = 0.007), but none within humidity and temperature data (Fig. S3, Table S9). Pearson’s product-moment correlation analysis showed no significant correlation between 5-Year Journal Impact Factors and Hedge’s g or reported p-values, even after removing outliers among Hedge’s g and p-values (Fig. S2). Furthermore, p-values extracted from the studies were examined to check for potential evidence of “p-hacking”. Visual inspection of the distribution of p-values surrounding p = 0.05 found limited evidence of p-hacking, with a small spike in the number of studies with p-values just under p = 0.05 (Fig. S4).

## Discussion

The COVID-19 pandemic has underscored the critical importance of understanding the risk factors that precipitate animal-borne disease outbreaks. While previous quantitative syntheses have identified general patterns associated with major drivers of zoonotic disease risk (5, 12), they often lack granular insights into specific climate impacts across various zoonotic diseases, highlighting the need for more detailed examinations of climate’s role in zoonotic disease dynamics. Our study provides evidence that climate sensitivity is widespread across a diverse range of zoonotic disease systems, suggesting that climate change is likely to further impact these systems in complex ways. However, significant biases and methodological gaps in current research hinder our ability to predict specific mechanisms, locations, and magnitudes of these impacts. By identifying the variability and limitations in our knowledge, we highlight the urgent need for standardised and transdisciplinary approaches to better predict how, where and through which mechanisms climate change will affect the global burden of zoonotic diseases.

The clearest result from our analysis is the positive effect of temperature on zoonotic disease risk for diseases that include arthropod vectors in the transmission cycle. This reflects findings from earlier studies, which have documented links between climate change and the spread of mosquito– and tick–borne diseases such as Dengue, Lyme, and West Nile virus into novel regions (13). Moreover, we found that nearly all study sites with Positive temperature effect sizes are located in regions projected to experience significant temperature increases exceeding 1.5°C, with a substantial proportion of sites also surpassing the 2°C threshold. This alignment suggests that these regions may face amplified risks under future climate scenarios.

In contrast, we did not find any consistent evidence that precipitation and humidity exert strong, uniform effects on zoonotic disease risk. The distribution of effect sizes among precipitation climate sensitivity data suggests that precipitation may drive large effects in zoonotic disease risk more frequently, possibly via the provisioning of resources (e.g. masting events affecting rodent populations and Puumala virus transmission) (32, 33). Notably, many sites with Positive precipitation effect sizes are projected to experience substantial rainfall increases, suggesting that these regions may see elevated disease risks in certain contexts. However, without more consistent and representative data this relationship remains uncertain. The considerable variation in reported effects across different diseases suggests that the influence of these climatic factors may be highly context-dependent and could vary with local ecological and socio-economic conditions.

There are numerous biases and limitations across the zoonotic disease literature that hinder our ability to generalise findings. The analysed studies predominantly focus on rodent-borne diseases, with bat-associated diseases notably underrepresented despite their key role in recent high-impact spillover events like Ebola and COVID-19, and the general uptick in epidemiological studies examining bat-borne diseases since 2003 (11). We identified studies on only 51 diseases, which represents just ∼6% of the (at least) 816 known human zoonotic pathogens (34). This taxonomic and geographic bias highlights the need for a more comprehensive and globally representative approach to studying the climate sensitivity of zoonotic diseases.

Methodological limitations further obstruct our understanding. Over half of the studies (57%) did not report a clear sample size used to calculate reported statistics, making it difficult to assess the reliability of their results. Although we identified over 50 distinct statistical methods reported, most studies either did not use the methods, or did not clearly state they used them, that can control for ubiquitous biases often seen in epidemiological data, such as spatial and temporal autocorrelation, and detection and reporting biases. Therefore, undertaking a formal meta-analysis to estimate an unbiased summary of the relationships we report is challenging.

Additionally, there was a notable lack of biological justification for the models used. Both theoretical (22) and empirical evidence (23, 27, 35, 36) suggests that relationships between climate and pathogen transmission are often complex and nonlinear, depending on the thermal biology of the vectors and pathogens involved. Yet, only 12% of the studies investigated these nonlinear effects. For instance, many vectors have optimal temperature ranges for transmission, and exceeding these optima can reduce transmission efficiency (22). As the direction and magnitude of climate effects on disease risk can depend heavily on the specific temperature ranges studied and the local environmental context, ignoring these nonlinear dynamics may lead to misleading inferences about climate impacts.

Overall, our study underscores the substantial heterogeneity in climate sensitivity across zoonotic diseases, as evidenced by the wide variation in effect sizes—even within specific pathogens (as illustrated in Figure 3). This heterogeneity across both vectored and non-vectored diseases complicates our ability to generalise findings and draw consistent conclusions about the impacts of climatic factors on zoonotic disease risk. However, it also highlights that climate sensitivity is a widespread phenomenon affecting a broad spectrum of zoonotic diseases, not just those transmitted by vectors. It remains unclear whether this variation represents true differences in disease responses to climate across different contexts or is an artefact of methodological inconsistencies and geographic sampling biases. This ambiguity highlights a critical gap in our understanding.

Given these challenges, and considering that many climate-sensitive diseases are located in areas consistently projected to experience significant increases in temperature—and in some cases, precipitation— there is a pressing need for more systematic and standardised research approaches. Adopting consistent methodologies that account for nonlinear relationships and control for common epidemiological biases will be essential. Such approaches would enable us to distinguish genuine ecological patterns from methodological noise, improving our ability to predict the mechanisms, locations, and magnitudes of climate change impacts on zoonotic diseases. Enhancing the rigour and transparency of future studies is crucial for informing effective public health interventions and policy decisions in the face of a changing climate.

## Methods

### Systematic review of literature

We conducted a systematic literature search to identify peer-reviewed quantitative studies addressing the effect of climatic factors (i.e., temperature, precipitation, and humidity) on components contributing to overall zoonotic burden. The databases searched included PubMed (to narrowly capture healthcare related studies) and Google Scholar (to ensure wide coverage of disciplines and journals), and the search was conducted between 15th - 22nd November 2022. To ensure thoroughness, a list of 51 generic zoonotic disease names and their synonyms were compiled (see Table S2). Terms describing zoonotic burden/risk were chosen to encompass all potential stages where disease risk could be influenced (i.e., hazard, exposure, and vulnerability). Searches were structured by pairing a specific disease (e.g., Brucellosis) with “Climat*” as subject headings. “Climat* incorporated alternative keywords such as “Climate change”, “Climatic change”, “Climate sensitive” or “Climate conditions”. To refine searches, Boolean operators were used to apply additional parameters (abundance/incidence/seroprevalence/cases) to search queries. Detailed search structures and example strings of search terms are provided in Table S1.

### Inclusion criteria and screening

The inclusion criteria for this study were quantitative field studies directly evaluating correlations between climatic factors and metrics of zoonotic risk. The climatic parameters were defined as temperature, precipitation, and humidity. Eligible studies were identified through a 3-step screening process: first the titles, then the abstracts followed by the full-texts were reviewed for relevance (see Supplementary File 1). For each search query, the first 50 returned titles were reviewed against the inclusion criteria. As eight searches were performed for each disease (Supplementary File 1), reviewing the top 50 titles per search gathered an exhaustive list of titles while reducing literature screening time. Following the removal of duplicate entries, abstracts were further reviewed for relevance to the research question. Finally, a full-text review of relevant articles was completed to extract data on climatic factors and zoonotic burden/risk.

### Data extraction and effect size calculations

Statistical values, associated sample sizes, and when available, dispersion values (e.g., standard error, standard deviation, 95% confidence intervals) were extracted from compiled articles. We also extracted geographical data including continent, country, locality, longitude and latitude of study sites. Data presented in text or tables was directly extracted and data from figures was digitised using WebPlotDigitizer (37). Data quality was assured by confirming interpretations (i.e., increases or decreases in zoonotic risk) with the studies own assessment of described trends.

For this investigation we defined a standardised measure of effect size using Hedge’s g (38). Observations were converted from presented statistics (i.e., odds ratio, relative risk, regression coefficient, correlation coefficient, t-, z-, f-, χ2 statistics) to Hedge’s g using standard conversion equations within the {esc} R package (39). The direction of the Hedges g value (i.e. a Positive or Negative value) determined whether a value represented an increase or decrease in zoonotic risk. To validate the robustness of conversion techniques, unit testing was performed using {tinytest} (40) and {testthat} packages (41) in R. Additionally, linear regressions were performed using the *lm* function from the {stats} package to measure the strength of correlations between obtained effect sizes and original statistical values (see Fig. S5).

Each study was categorised by the disease studied (e.g. Hantaviral diseases, Echinococcoses, Arenaviral diseases), risk/hazard metric (e.g. host disease incidence, host infection, host abundance), principal reservoir of the disease (e.g. rodents, mammals, livestock), pathogen type causing the disease (Virus, Bacteria, Parasite), type of the relationship between climate and disease risk (linear, non-linear) and modelling method used (e.g. GL(M)M, Correlation, (S)ARIMA) (Table S1, Fig. 1). Additionally, diseases were categorised based on their mode of transmission into two groups: vectored and non-vectored diseases.

### Statistical evaluation of reported direction statements

We used Pearson’s Chi-squared Test for Count Data (from the {stats} R package) to investigate if the proportions of reported increases and decreases in disease risk were significantly different. We performed bootstrap resampling with 1000 iterations, sampling 80% of the data with replacement in each iteration. The 80% resampling was chosen to balance between reducing variance and maintaining sufficient sample size for reliable estimation. Specifically, we performed Chi-squared tests on each bootstrap sample for the entire dataset and for the three climate subsets: temperature, precipitation and humidity.

To ensure robustness, we repeated the procedure by sequentially removing records from one major group at a time (e.g. separately removing records from each of the top three diseases) from the analysis, to identify which categories significantly affected the proportions of increases to decreases in disease risk. The groups removed in this process and the percentage of the analysed dataset these groups represent included: Hantaviral diseases (24.5%), Arboviral diseases (22.5%), Leptospirosis (11.4%) (top three diseases); China (33.7%), Iran (8.7%), USA (6.4%) (top three countries); rodents (39.1%), mammals - multispecies (21.8%), livestock (16.8%) (top three principle reservoirs); viruses (56.4%), bacteria (37.9%), parasites (5.7%) (top three pathogen types); Pearson correlation (12.1%), Spearman rank correlation (11.1%), negative binomial regression (3.5%) (top three statistical methods).

### Analysis of standardised effect sizes

We categorised effect sizes into three groups based on the value of Hedge’s g: Positive effect (g > 0.2), No effect (-0.2 < g < 0.2) and Negative effect (g < -0.2). These thresholds were chosen based on Cohen’s conventions for small effect sizes (42), where values of 0.2 represent a small effect. Additionally, we analysed the association between the reported direction of the climate impacts on disease risk (Increase/Decrease/No change) and the effect size categories based on our Hedge’s g calculations (Negative effect/No effect/Positive effect). To robustly estimate the significance of this association, we performed a bootstrap analysis with 1000 samples. Each bootstrap sample consisted of 80% of the original data, sampled with replacement. We applied Fisher’s exact test to each bootstrap sample to determine the p-value for the association between the two variables.

To assess whether climate effects on measures of disease risk are uniform across different subsets of the data (specific transmission types, reservoirs, pathogens, vectors, countries and diseases), we compared the effect size distribution for each subset to the full effect size distribution for each climate factor (temperature, precipitation, and humidity). We performed a two-sample permutation-based Anderson-Darling test with 1000 replicates between the subset effect size distribution and the full distribution to test whether they are likely to be from the same distribution. The Anderson-Darling test was chosen because it can be used as a powerful non-parametric test that is sensitive to differences in both the location and shape of distributions, especially in the tails. As one distribution is a subset of the other, we also performed the test removing the subset from the full distribution.

We additionally compared the effect size distributions for vectored and non-vectored diseases using Kolmogorov-Smirnov test to investigate whether the climate factors show the same effect between these groups.

### Analysis of projected climatology data across study sites

To assess the co-occurrence of climate-sensitive diseases with areas expected to experience significant shifts in climatic conditions with future climate change, we extracted modelled climate shifts for study sites locations that reported Positive (Hedge’s g > 0.2) or Negative (Hedge’s g < -0.2) climate sensitivity effects.

To determine expected climate shifts, we used bioclimatic data from the CHELSA V2.1 CMIP6 dataset (43, 44), focusing on two climate variables: BIO1 (mean annual air temperature) and BIO12 (annual precipitation amount). These variables were extracted for both the baseline period (1981–2010) and a future time window (2041–2070) under three Shared Socioeconomic Pathways (SSP1-RCP2.6, SSP3-RCP7.0, SSP5-RCP8.5). We selected five GCMs (Global Climate Models) (GFDL-ESM4, IPSL-CM6A-LR, MPI-ESM1-2-HR, MRI-ESM2-0, UKESM1-0-LL) to capture variability across model predictions and to ensure a robust comparison across different climate scenarios.

For each spatial point corresponding to a study location, we extracted baseline and future BIO1 and BIO12 values from the climate rasters. This allowed us to compute the expected change in temperature (°C) and precipitation (mm) between the baseline and future projections. The differences were calculated for each combination of SSP and GCM, yielding 15 projected differences for each site to account for the variability and uncertainty inherent in climate projections and emission scenarios.

To capture the magnitude of climate shifts at each location, we categorised the temperature and precipitation differences based on a set of three temperature and three precipitation thresholds. For temperature, for each spatial point, we identified the proportion of SSP x GCM combinations predicting increases greater or less than 1°C, 1.5°C, and 2°C. Similarly, for precipitation, we calculated the proportion of SSP x GCM combinations predicting increases or decreases beyond ±25 mm, ±50 mm, ±100 mm, and the proportions of combinations falling in the intermediate range (e.g. proportion of predictions > + 25mm, between - 25mm to + 25mm and < -25mm). Then, for each of the three temperature and precipitation thresholds we summarised the modal categories of climate change for each location across the 15 SSP x GCM combinations, as there was generally strong agreement about the direction and magnitude of expected shifts.

Transformation of continuous climate data into a set of discrete categories of temperature and precipitation changes allowed us to assess whether some locations with Positive (Hedge’s g > 0.2) or Negative (Hedge’s g < -0.2) climate sensitivity effects are associated with a certain magnitude or direction of climate change. We constructed six contingency tables (for three temperature and three precipitation thresholds, see Fig. S6) and compared the association between disease climate sensitivity effects (Positive/Negative Hedge’s g) and the magnitude of predicted climate changes. Depending on the structure of the contingency table, either Pearson’s Chi-squared Test for Count Data (if all cells in the contingency table had 5 or more counts) or Fisher’s Exact Test for Count Data was applied to evaluate the association between these sensitivity categories and climate change magnitudes. We performed the tests on 1000 bootstrap replicates for each contingency table, and calculated mean p-values, test statistics, 95% confidence intervals and also the percentage of iterations with a significant (p < 0.05) test result.

### Publication bias assessment

To assess publication bias, we investigated the correlation between the 5-Year Impact Factors of journals included in our study and their respective Hedge’s g and p-values. We selected Pearson’s product-moment correlation test (from {stats} package) to evaluate the correlations. Additionally, we applied the Interquartile Range (IQR) method to detect the outliers within the Hedge’s g and p-value data. Outliers were defined as values that fell more than 1.5 times the IQR below the first quartile or above the third quartile. The Identified outliers included 66 of Hedge’s g values and 30 of p-values. To examine whether these outliers influenced the direction or significance of the correlations, we performed Pearson’s product-moment correlation tests twice: before and after removing the outliers (Fig. S2).

Furthermore, we prepared funnel plots visualising the distribution of effect sizes against their standard errors, where such calculations were possible. The funnel plots were created for the overall dataset, and for the three climate factors (temperature, precipitation and humidity). To each plot, we applied Egger’s regression test from the {metafor} package (45) to check for significant asymmetry, indicating potential publication bias. Egger’s test assesses whether there is a linear relationship between the effect sizes and their standard errors; a significant result suggests asymmetry in the funnel plot, which may be due to publication bias. To further investigate potential “p-hacking,” we visually inspected the distribution of p-values reported in the studies, focusing on the values surrounding p = 0.05.

### Analysis environment

All parts of the analysis were conducted using R version 4.3.1. The full dataset and R code used to perform the analyses and create the figures is available at: https://github.com/BioDivHealth/climate_meta.

## Data Availability

All data produced in the present study are available upon reasonable request to the authors

https://github.com/BioDivHealth/climate_meta

## Acknowledgements

This research was supported by the UKRI Biotechnology and Biological Sciences Research Council (BBSRC) grants (Refs: BB/X005364/1 and BB/X005364/2), the Wellcome Trust grants (Refs: 220179/Z/20/Z, 220179/A/20/Z, and 226080/Z/22/Z), the National Science Foundation (NSF) grant (Ref: 2213854), and the Wolfson Foundation (via a UCL Excellence Fellowship; RG).

## Author contributions

D.W.R., L.G., N.I. and R.G. designed research, L.G., A.T. R.G. and D.W.R. performed research, L.G. and A.T. collected data, A.T., L.G., R.G. and N.I. analysed data, A.T., L.G., N.I., D.W.R and R.G. wrote the paper.

## Supplementary Figures

**Figure S1.**
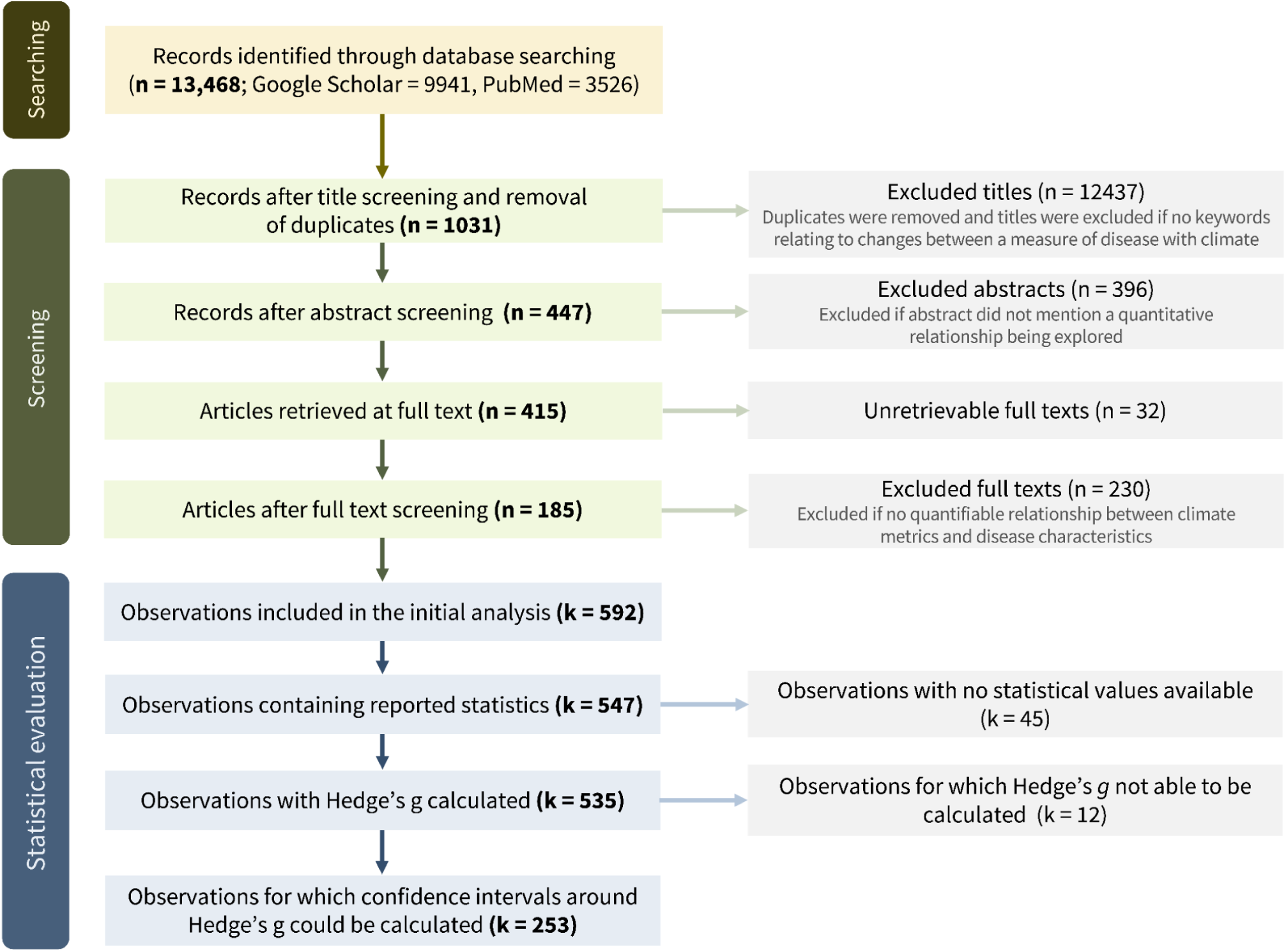
Flow chart visualising the process of literature search, literature screening and statistical evaluation of extracted data. N values correspond to the number of scientific articles and k values correspond to the number of observations extracted.

**Figure S2.**
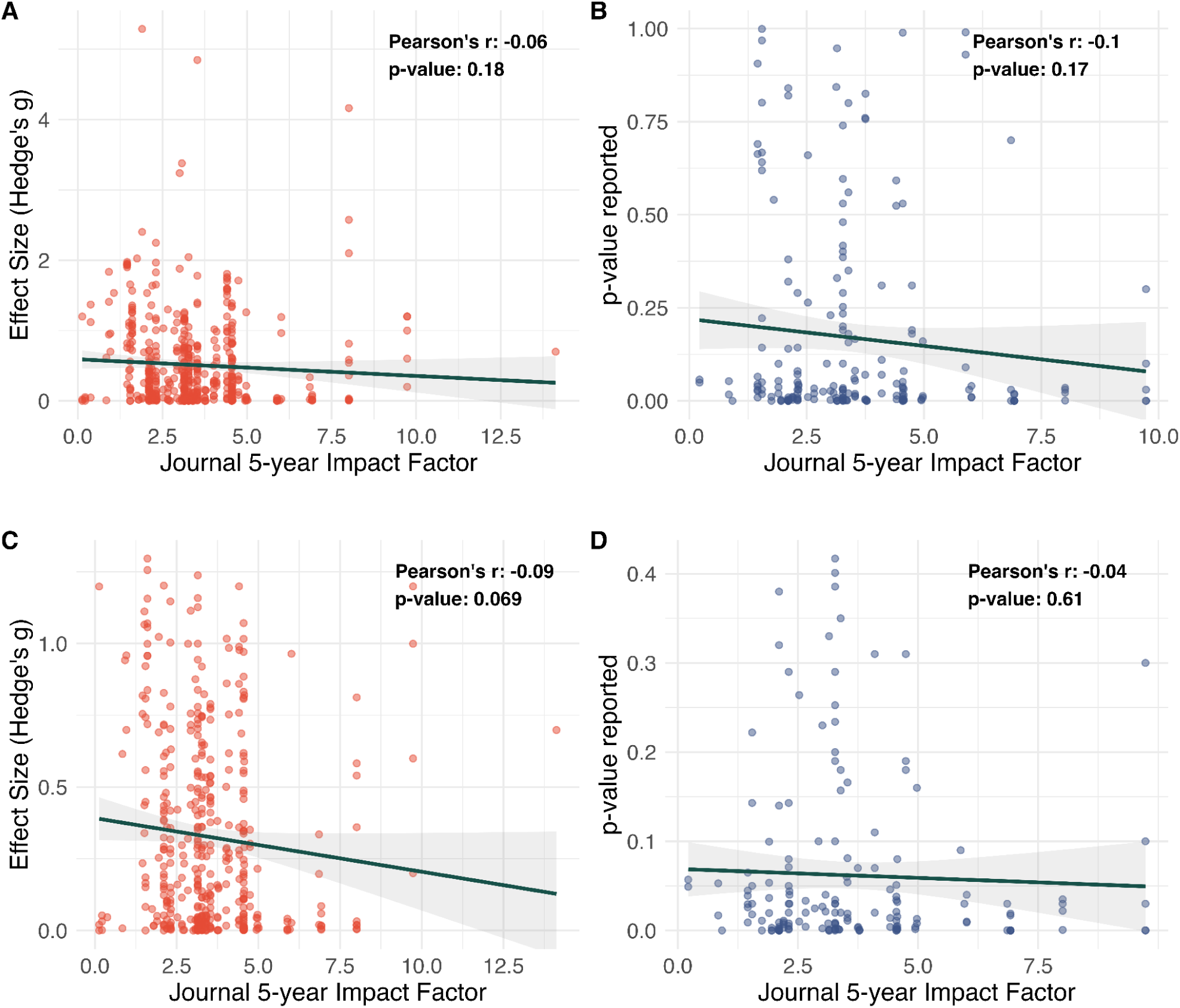
Hedge’s g effect sizes (A,C) and reported p-values (B,D) plotted against 5-year Impact Factors of journals in which corresponding studies were published. Results of Pearson’s product-moment correlation test (correlation coefficient and p-value) are annotated in the upper right corner of each subplot. Subplots A and B are based on full dataset and subplots C and D were made following removal of outliers among Hedge’s g and p-value data using IQR method.

**Figure S3.**
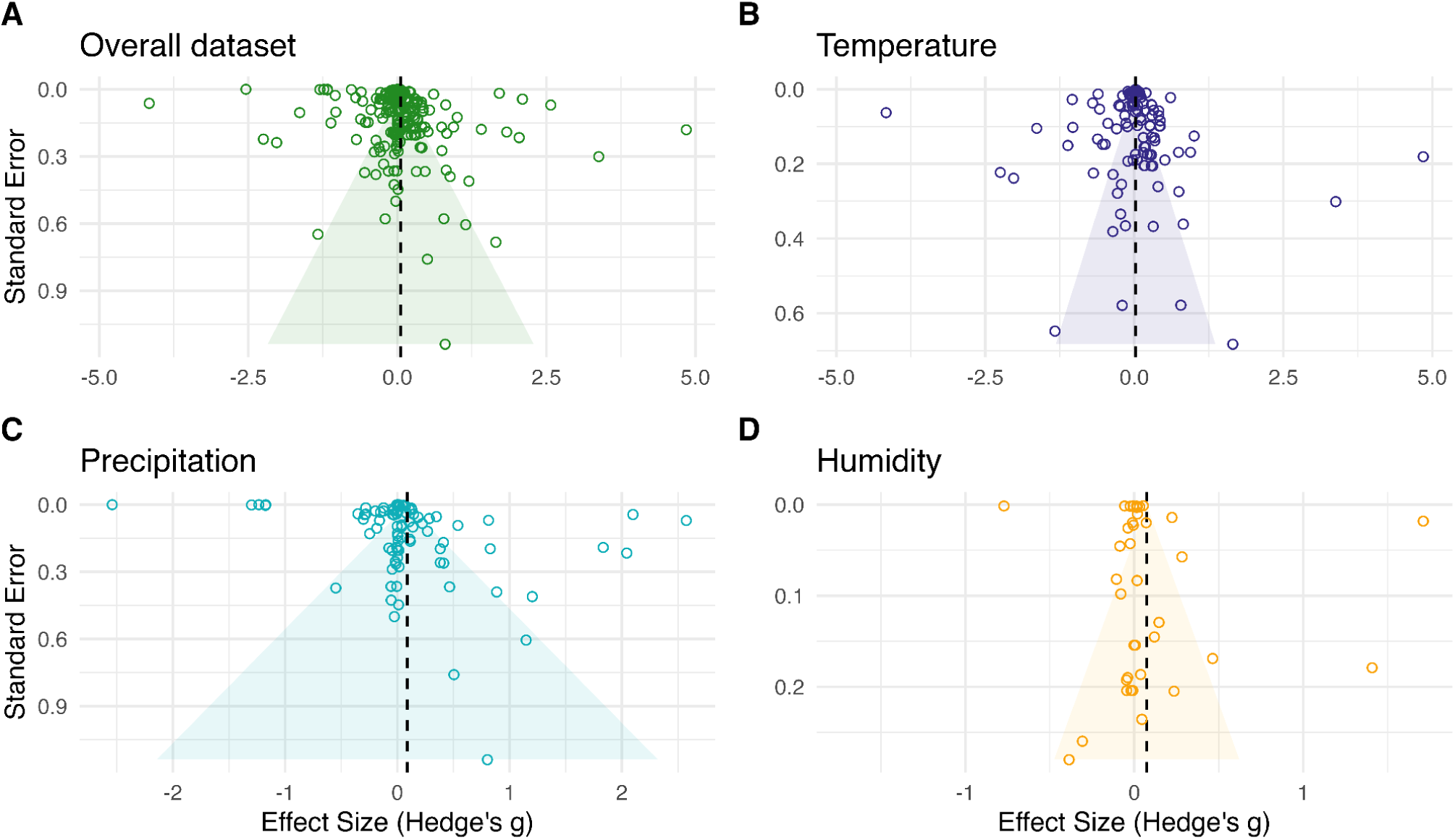
Funnel plots of standard errors against corresponding Hedge’s g values. Plot **A** made for the overall dataset, plots **B**, **C** and **D** made for Temperature, Precipitation and Humidity data respectively. Vertical dotted line represents the mean effect size within each plot. Shaded area represents 95% confidence region constructed for each plot.

**Figure S4.**
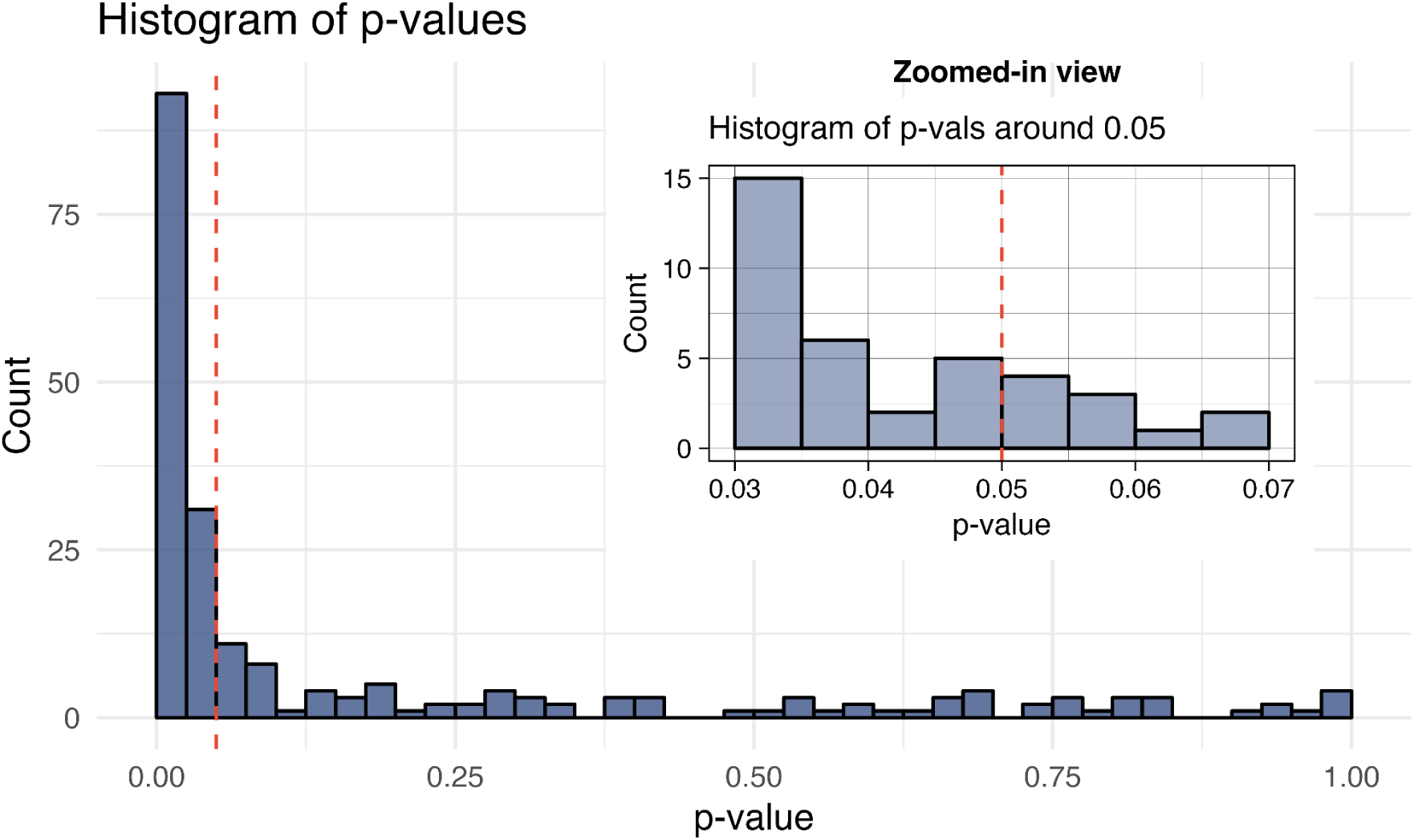
Distribution of p-values in study sample. The main plot shows the overall distribution of p-values from all studies for which numeric p-values were provided. X-axis represents the p-values, and Y-axis represents the count of p-values falling within a specific bin. The inset plot shows the distribution of p-values where 0.03 < p < 0.07. Red vertical lines signify p-value = 0.05.

**Figure S5.**
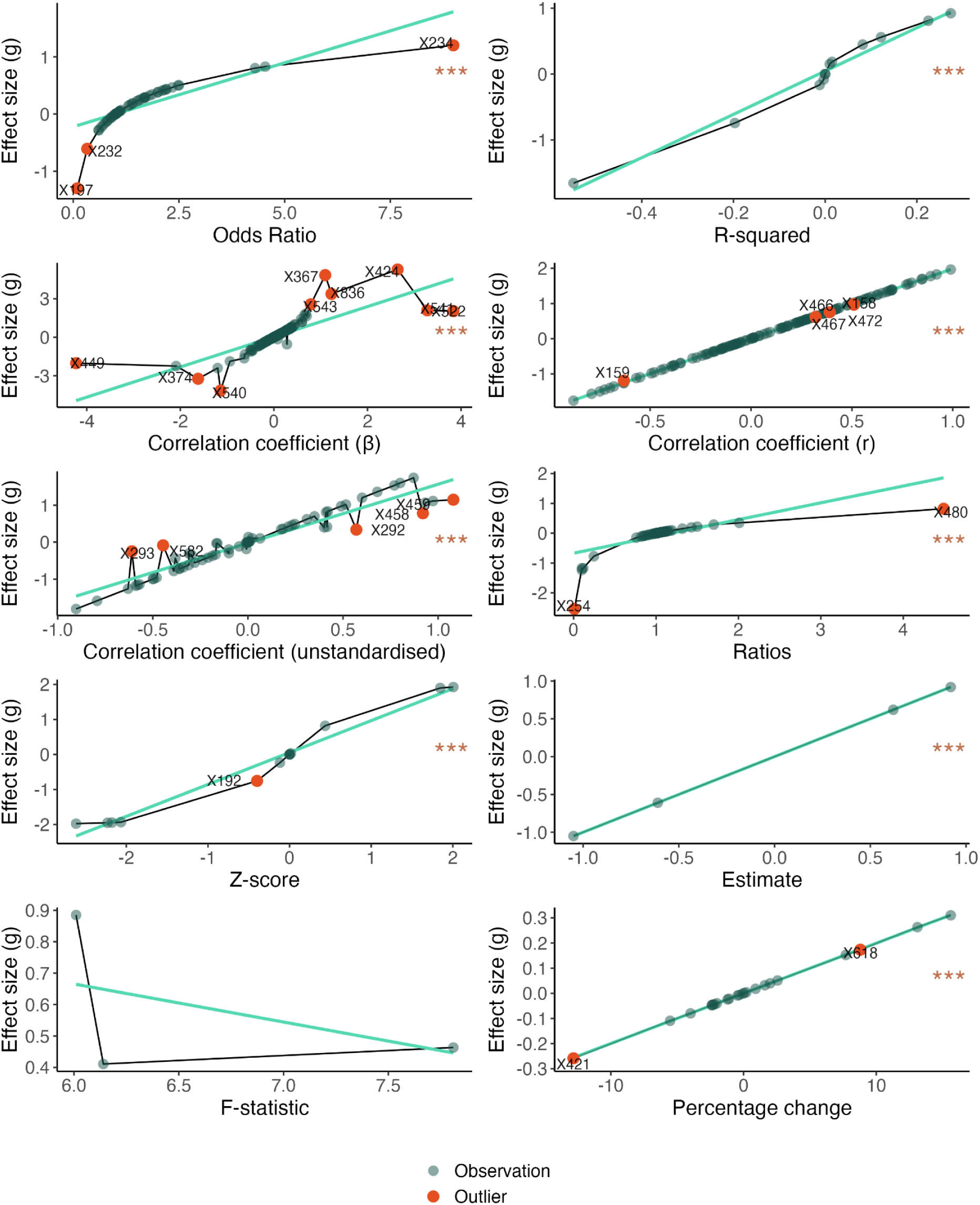
Relationships between extracted statistical measures and corresponding Hedge’s g values. The plots show the relationship between different statistical measures (e.g., Odds Ratio, Correlation Coefficient, F-statistic) sourced from studies and the calculated effect sizes (Hedge’s g). Dark green points indicate individual observations and red points highlight outliers. Outliers were determined based on residuals from a linear regression model, where points with residuals greater than two standard deviations from the mean were marked as outliers. The light green line represents the linear fit for each measure.

**Figure S6.**
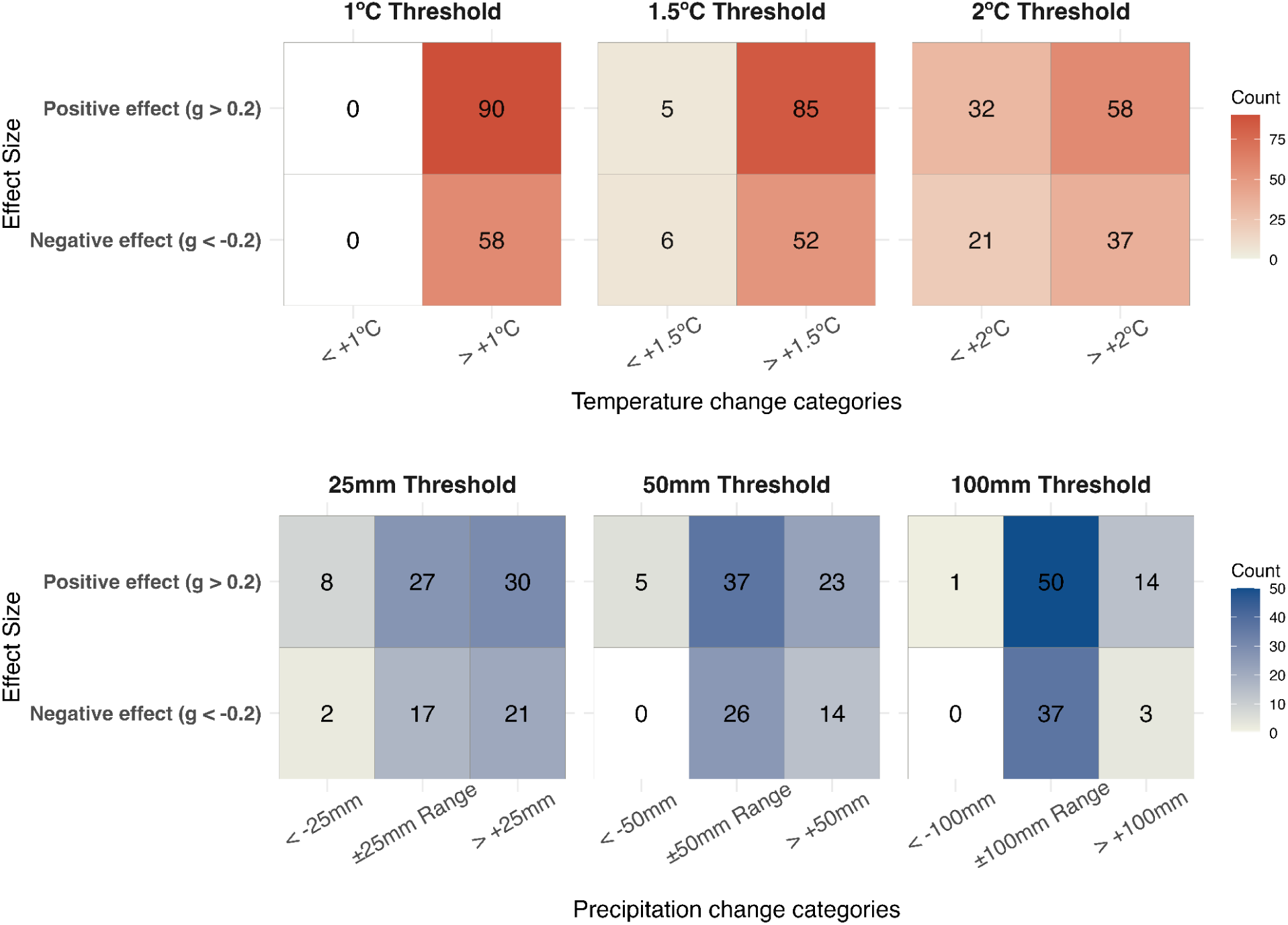
Contingency tables showing the association between disease climate sensitivity effect sizes (Hedge’s g) and predicted changes in temperature and precipitation across study locations, based on future climate projections from the CHELSA V2.1 CMIP6 dataset. The top row represents temperature thresholds of 1°C, 1.5°C, and 2°C, while the bottom row represents precipitation thresholds of ±25mm, ±50mm, and ±100mm. Positive (Hedge’s g > 0.2) and Negative (Hedge’s g < -0.2) climate sensitivity effect sizes are shown in relation to predicted climate change categories. The numbers in each tile represent the count of locations where the most frequent (modal) category of climate change, determined from 15 projections (comprising five Global Climate Models and three Shared Socioeconomic Pathways), was associated with the respective sensitivity effect size.

## Supplementary Tables

**Table S1.**
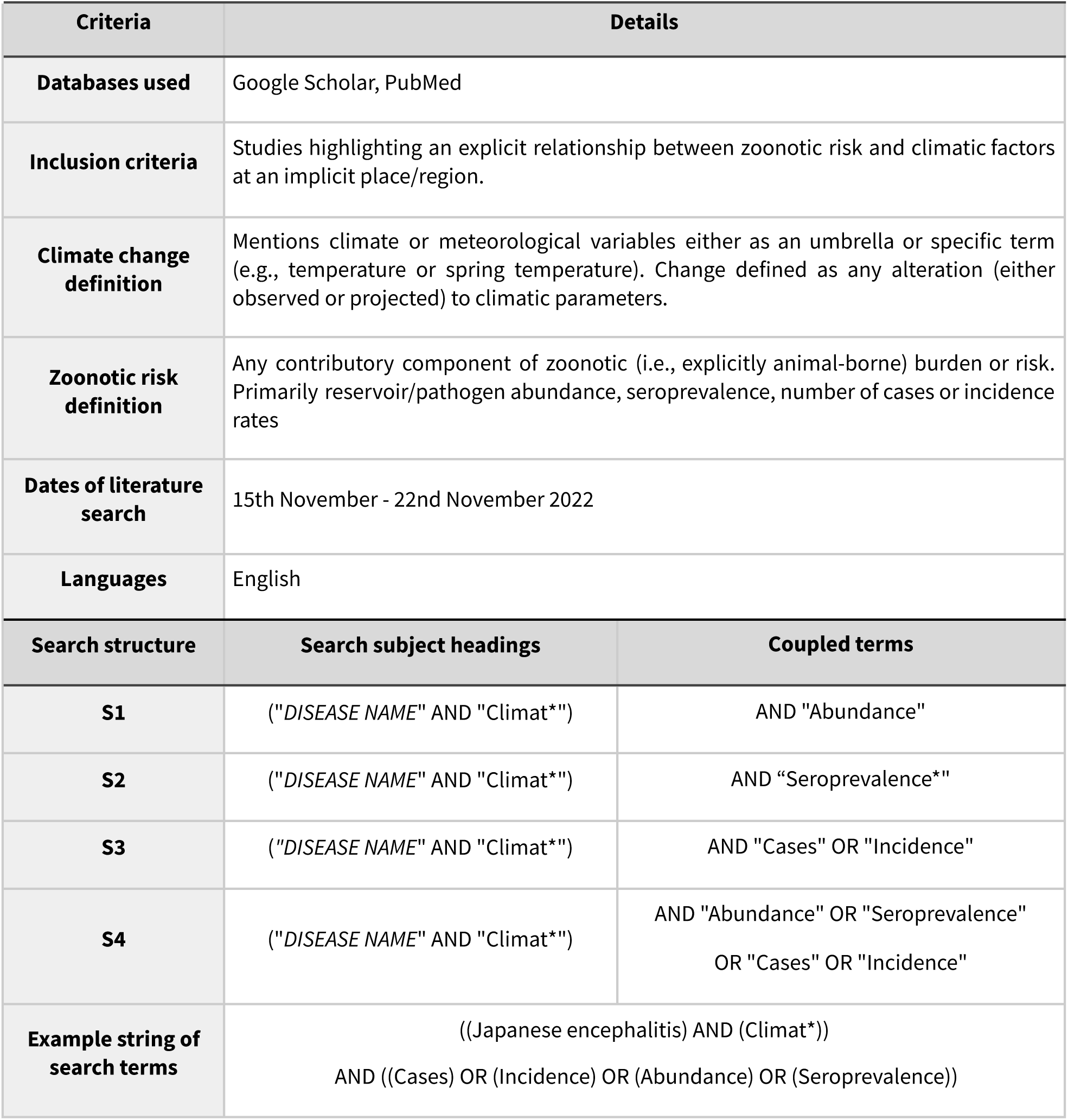
Table of the inclusion criteria, definitions and search structures used to identify relevant articles during systematic review. “DISEASE NAME” refers to the specific disease targeted by each search.

| Criteria | Details |  |
| --- | --- | --- |
| **Databases used** | Google Scholar, PubMed |  |
| **Inclusion criteria** | Studies highlighting an explicit relationship between zoonotic risk and climatic factors at an implicit place/region. |  |
| **Climate change definition** | Mentions climate or meteorological variables either as an umbrella or specific term (e.g., temperature or spring temperature). Change defined as any alteration (either observed or projected) to climatic parameters. |  |
| **Zoonotic risk definition** | Any contributory component of zoonotic (i.e., explicitly animal-borne) burden or risk. Primarily reservoir/pathogen abundance, seroprevalence, number of cases or incidence rates |  |
| **Dates of literature search** | 15th November - 22nd November 2022 |  |
| **Languages** | English |  |
| Search structure | Search subject headings | Coupled terms |
| **S1** | ("DISEASE NAME" AND "Climat*") | AND "Abundance" |
| **S2** | ("DISEASE NAME" AND "Climat*") | AND "Seroprevalence" |
| **S3** | ("DISEASE NAME" AND "Climat*") | AND "Cases" OR "Incidence" |
| **S4** | ("DISEASE NAME" AND "Climat*") | AND "Abundance" OR "Seroprevalence"<br>OR "Cases" OR "Incidence" |
| **Example string of search terms** | ((Japanese encephalitis) AND (Climat*))<br>AND ((Cases) OR (Incidence) OR (Abundance) OR (Seroprevalence)) |  |

**Table S2.** Names of zoonotic diseases used in literature search. Each name replaced the “DISEASE NAME” field in the string of search terms presented in Table S1. In some cases, additional synonyms of the disease names were used, these can be found in Supplementary File 1, along with complete search structures.

| Disease name |  |  |
| --- | --- | --- |
| Alveolar echinococcosis | Japanese encephalitis | Flinders island spotted fever |
| Leptospirosis | Crimean-Congo hemorrhagic fever | Japanese spotted fever |
| Monkeypox | Penicilliosis | American tick-bite fever |
| Ohio valley disease | Boutonneuse fever | Sealpox |
| Brucellosis | Scrub typhus | Lymphocytic choriomeningitis |
| Leprosy | Psittacosis | Giardiasis |
| Hemorrhagic fever | Rickettsiosis | Zoonotic tuberculosis |
| Rabies | Rocky mountain spotted fever | Rift valley fever |
| Lassa fever | Menangle virus | Fascioliasis |
| Nephropathia epidemica | Venezuelan hemorrhagic fever | Anthrax |
| Hantaviral Disease | Bolivian hemorrhagic fever | Zoonotic Influenza |
| Hantavirus Pulmonary Syndrome | Mediterranean spotted fever | Plague |
| Nipah virus | African tick-bite fever | Argentine Hemorrhagic fever |
| Hendra virus | Tacaribe virus | Cowpox |
| Paracoccidioidomycosis | Raccoon roundworm infection | Marburg virus |
| Ebola | Siberian tick typhus | Encephalitis |
| Severe Acute Respiratory Syndrome | Queensland tick typhus | Venezuelan equine encephalitis |

**Table S3.**
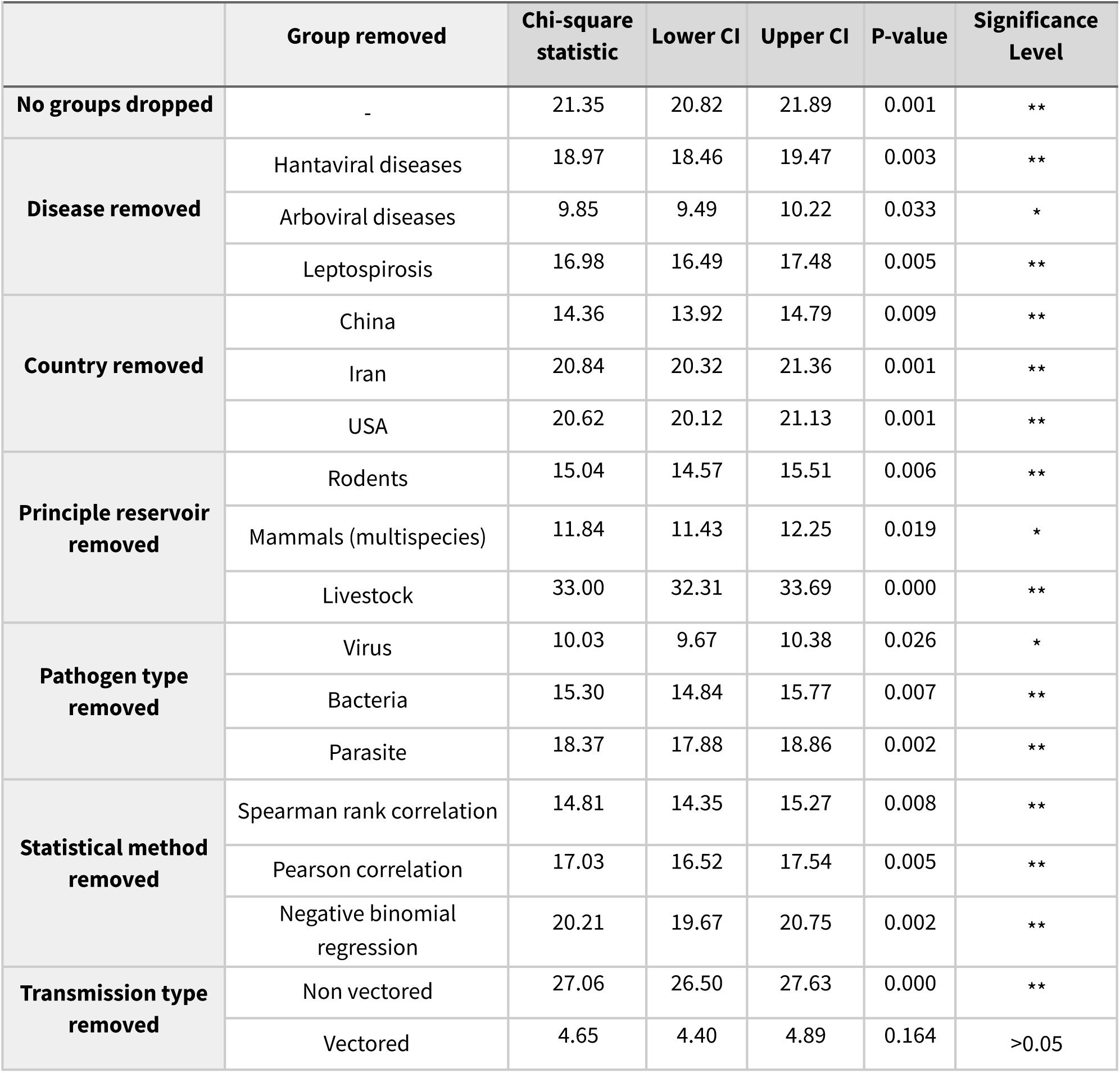
Results of Pearson’s Chi-squared Test for Count Data conducted on the data describing reported direction (increase vs decrease) of the impacts of all climate effects on disease risk for the overall dataset. The chi-squared goodness-of-fit tests assessed whether the observed frequencies of reported increases and decreases in disease risk significantly differed from the expected frequencies under the null hypothesis (i.e., no difference in the likelihood of increases and decreases). First row shows results of chi-square tests using the full dataset. Subsequent rows report the results of tests in which major diseases, countries, reservoirs, pathogen types, statistical methods and transmission types were removed from the analyses one-at-a-time. Significant results suggest an uneven proportion of increases to decreases of disease risk reported. Chi-square tests were run on 1000 bootstraps of 80% of the data. * = p < 0.05; ** = p < 0.01.

**Table S4.**
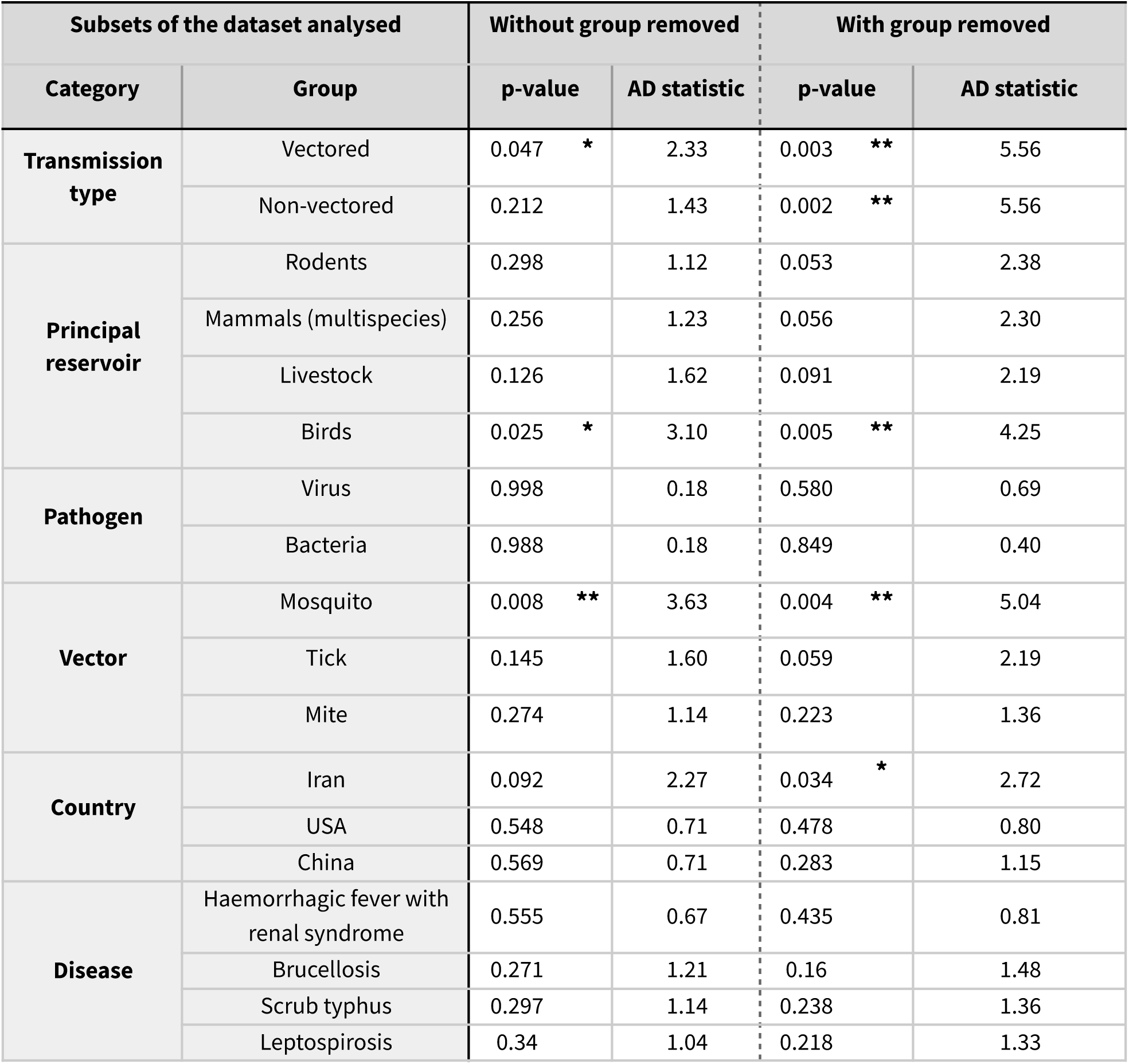
Results of Permutation-based Anderson-Darling test of homogeneity evaluating whether the distribution of effect sizes within specific groups (e.g., transmission types, principal reservoirs, pathogen types, or vector types) significantly differs from the overall distribution of effect sizes for temperature. The table compares the effect size distributions within a given group to the full temperature dataset (“Without group removed”) and to the full temperature dataset with that specific group excluded (“With group removed”). p-values and AD statistics are provided for both comparisons, with asterisks indicating significance levels (* = p < 0.05; ** = p < 0.01).

**Table S5.** Results of the Exact two-sample Kolmogorov-Smirnov Tests comparing the distribution of Effect Sizes (Hedge’s g) between Vectored and Non-vectored diseases for different environmental conditions. The table included the test statistics and corresponding p-values for differences in effect size distributions between vectored and non-vectored transmission types under three environmental conditions: Temperature, Precipitation, and Humidity. A significant p-value suggests a difference between the distributions of the two groups. The test statistic represents the maximum difference between the empirical cumulative distribution functions (ECDFs) of the two groups.

| Variable | p-value | D^+ Test statistic |
| --- | --- | --- |
| Temperature | 0.001 ** | 0.26 |
| Precipitation | 0.445 | 0.09 |
| Humidity | 0.079 | 0.23 |

**Table S6.** Results of Permutation-based Anderson-Darling test of homogeneity evaluating whether the distribution of effect sizes within specific groups (e.g., transmission types, principal reservoirs, pathogen types, or vector types) significantly differs from the overall distribution of effect sizes for humidity. The table compares the effect size distributions within a given group to the full humidity dataset (“Without group removed”) and to the full humidity dataset with that specific group excluded (“With group removed”). p-values and AD statistics are provided for both comparisons, with asterisks indicating significance levels (* = p < 0.05; ** = p < 0.01).

| Subsets of the dataset analysed |  | Without group removed |  | With group removed |  |
| --- | --- | --- | --- | --- | --- |
| Category | Group | p-value | AD statistic | p-value | AD statistic |
| Transmission type | Vectored | 0.898 | 0.35 | 0.427 | 0.88 |
|  | Non-vectored | 0.977 | 0.24 | 0.442 | 0.88 |
| Principal reservoir | Rodents | 0.796 | 0.44 | 0.432 | 0.88 |
|  | Mammals (multispecies) | 0.4 | 0.89 | 0.127 | 1.64 |
|  | Livestock | 0.036 * | 2.74 | 0.002 ** | 4.75 |
|  | Birds | 0.731 | 0.52 | 0.651 | 0.59 |
| Pathogen | Virus | 0.964 | 0.26 | 0.662 | 0.61 |
|  | Bacteria | 0.966 | 0.24 | 0.433 | 0.83 |
| Vector | Mosquito | 0.444 | 0.92 | 0.273 | 1.20 |
|  | Tick | 0.586 | 0.67 | 0.403 | 0.86 |
|  | Mite | 0.754 | 0.50 | 0.605 | 0.68 |
| Country | China | 0.248 | 1.25 | 0.051 | 2.48 |
|  | Iran | 0.016 * | 3.43 | 0.002 ** | 5.02 |
|  | India | 0.958 | 0.28 | 0.923 | 0.32 |
| Disease | Haemorrhagic fever with renal syndrome | 0.234 | 1.20 | 0.138 | 1.62 |
|  | Brucellosis | 0.078 | 2.20 | 0.015 * | 3.32 |
|  | Scrub typhus | 0.742 | 0.50 | 0.589 | 0.68 |
|  | Leptospirosis | 0.762 | 0.50 | 0.551 | 0.70 |

**Table S7.** Results of Permutation-based Anderson-Darling test of homogeneity evaluating whether the distribution of effect sizes within specific groups (e.g., transmission types, principal reservoirs, pathogen types, or vector types) significantly differs from the overall distribution of effect sizes for precipitation. The table compares the effect size distributions within a given group to the full precipitation dataset (“Without group removed”) and to the full precipitation dataset with that specific group excluded (“With group removed”). p-values and AD statistics are provided for both comparisons, with asterisks indicating significance levels (* = p < 0.05; ** = p < 0.01).

| Subsets of the dataset analysed |  | Without group removed |  | With group removed |  |
| --- | --- | --- | --- | --- | --- |
| Category | Group | p-value | AD statistic | p-value | AD statistic |
| Transmission type | Vectored | 0.755 | 0.48 | 0.368 | 1.00 |
|  | Non-vectored | 0.981 | 0.22 | 0.358 | 1.00 |
| Principal reservoir | Rodents | 0.814 | 0.41 | 0.334 | 1.04 |
|  | Mammals (multispecies) | 1 | 0.10 | 0.999 | 0.17 |
|  | Livestock | 0.077 | 2.17 | 0.018 * | 3.02 |
|  | Birds | 0.237 | 1.27 | 0.125 | 1.72 |
| Pathogen | Virus | 0.568 | 0.69 | 0.096 | 2.03 |
|  | Bacteria | 0.723 | 0.54 | 0.207 | 1.40 |
| Vector | Mosquito | 0.455 | 0.83 | 0.345 | 1.11 |
|  | Tick | 0.392 | 0.94 | 0.315 | 1.09 |
|  | Mite | 0.477 | 0.85 | 0.385 | 1.00 |
| Country | China | 0.776 | 0.47 | 0.438 | 0.88 |
|  | Iran | 0.242 | 1.25 | 0.157 | 1.59 |
|  | Argentina | 0.241 | 1.27 | 0.222 | 1.32 |
| Disease | Haemorrhagic fever with renal syndrome | 0.494 | 0.80 | 0.374 | 0.96 |
|  | Brucellosis | 0.174 | 1.61 | 0.098 | 2.03 |
|  | Scrub typhus | 0.454 | 0.85 | 0.35 | 1.00 |
|  | Leptospirosis | 0.095 | 1.98 | 0.027 * | 2.83 |

**Table S8.** Results of Pearson’s Chi-squared Tests for Count Data and Fisher’s Exact Tests for Count Data conducted on the data describing associations between climate sensitivity (Positive/Negative Hedge’s g) and projected changes in temperature and precipitation across study sites. The tests were performed at three temperature thresholds (+1°C, +1.5°C, +2°C) and three precipitation thresholds (±25 mm, ±50 mm, ±100 mm) using predictions from three SSP scenarios and five GCMs for the period 2041–2070 compared to baseline conditions (1981–2010). The p-values, 95% confidence intervals around p-values, and test statistics are reported. Tests were run on 1000 bootstrap samples and the percentage of tests with p-value under 0.05 is reported.

| Variable | Threshold | Test | p-value | p-value<br>95% CI | Percentage of<br>p-values < 0.05 | Test statistic |
| --- | --- | --- | --- | --- | --- | --- |
| **Temperature** | +1°C | Chi-Square | 0.06 | 0.05 – 0.07 | 78% | 7.58 |
|  | +1.5°C | Fisher's Exact | 0.43 | 0.40 – 0.45 | 17% | – |
|  | +2°C | Chi-Square | 0.59 | 0.57 – 0.61 | 3% | 0.76 |
| **Precipitation** | ± 25mm | Fisher's Exact | 0.34 | 0.32 – 0.36 | 17% | – |
|  | ± 50mm | Fisher's Exact | 0.22 | 0.21 – 0.23 | 20% | – |
|  | ± 100mm | Fisher's Exact | 0.19 | 0.17 – 0.20 | 43% | – |

**Table S9.** Results of the Egger’s regression tests on funnel plot asymmetry. This table presents the results of Egger’s regression tests (***regtest {metafor}***) for funnel plot asymmetry, based on effect sizes and standard errors across the overall dataset and three climatic subsets: temperature, precipitation, and humidity. The z-value tests the null hypothesis of no funnel plot asymmetry, and the p-value indicates statistical significance. Significant p-values (p < 0.05) suggest publication bias.

| Data | Intercept | Z_value | P_value |
| --- | --- | --- | --- |
| Overall dataset | -0.054 | 2.622 | **0.009** |
| Temperature | -0.077 | 1.225 | 0.221 |
| Precipitation | -0.061 | 2.710 | **0.007** |
| Humidity | 0.073 | 0.050 | 0.960 |

## Supplementary Files

**Supplementary File 1.** Excel Worksheet containing sheets with: full dataset, metadata, studies included in the dataset, log of literature search. https://github.com/BioDivHealth/climate_meta/blob/main/data/SI_FILES/Supplementary_File_1.xlsx

